# Evaluation of Fecal Occult Blood Testing Kits for Rapid Point-of-Care Diagnosis of Invasive Diarrhea in Young Children

**DOI:** 10.1101/2023.02.01.23285337

**Authors:** David A. Kwasi, Pelumi D. Adewole, Olabisi C. Akinlabi, Stella E. Ekpo, Iruka N. Okeke

## Abstract

**Background:** Antimicrobials are only indicated in acute childhood diarrhea when the infection is invasive (and therefore often bloody), or persistent. Rapid and cost-effective screening for invasive diarrhea at the point of care can therefore inform treatment decisions. Multiple rapid fecal occult blood testing (FOBT) kit brands are widely available in Nigeria. This study aimed to compare the diagnostic utility of locally procurable FOBT kits in invasive infantile diarrhea to the innovators product, using fecal microscopy as the gold standard.

**Materials and Methods:** Fecal specimens from 46 children under 5 years old with diarrhea, being collected as part of ongoing case-control studies, were tested according to manufacturers’ instructions for each of five FOBT kits. Fecal microscopy for occult blood, and culture for bacterial pathogens were also performed concomitantly using standard procedures.

**Results:** Stool microscopy confirmed almost ubiquitous presence of white blood cells in stool from children with diarrhea, whereas red blood cells were less commonly detected. A positive FOBT reaction was only seen when red blood cells were present at more than trace levels and was partially correlated with the presence of a potentially invasive pathogen. Each of the five FOBT kits tested showed 54-61% sensitivity, 87-90% specificity, and acceptable positive- and negative-predictive values for invasive diarrhea.

**Conclusions:** Four inexpensive, locally available kits identified invasive pediatric diarrheas showed reasonable performance for detecting likely invasive diarrhea and performed similarly to the more difficult-to-procure innovator’s product. FOBT kits are a rapid option for presumptive diagnosis of invasive diarrhea, are a viable alternative to stool microscopy for paediatric specimens at the point-of-care, and could serve as early warning indicators for dysentery outbreaks.

## Introduction

Infectious acute diarrhea can be life -hreatening in children [1,2] and hence requires prompt diagnosis and management. Most diarrhea infections are self-resolving with rehydration being the principal intervention required [3]. The decision to use antimicrobials when indicated is best informed by stool microscopy, culture, and susceptibility tests. These tests require trained laboratory personnel and, in the case of culture, take a few days to complete [4]. Inevitably, initial therapeutic choices are not always the most appropriate, which can cause delays in instituting the right treatment or promote antimicrobial use when not indicated. A rapid and cost-effective screening process to identify likely invasive infections at the point-of-care is thus expedient. This is particularly true in Nigeria where diarrhea is an important cause of childhood illness and death and where antimicrobial overuse places the entire population at high risk of the consequences of antimicrobial resistance [5,6,7]. A range of conditions, including gastrointestinal cancers, malabsorption, abdominal pain, constipation and iron deficiency anemia can result in gross or occult blood in stool but the list of conditions includes shigellosis and other enteric invasive infections [8,9,10,11].

Blood in stool is ideally detected by microscopy but tests designed to detect haemoglobin are commonly used as a diagnostic aid for carcinomas and invasive diarrhea [12]. These spot tests do not require trained microscopists and are easy to perform. We observed that several brands of kits for these tests are available on the Nigerian market. Previously, Bardhan et. al. [13] investigated the role of clinical features, fecal microscopy (FM), and fecal occult blood testing (FOBT) in distinguishing invasive diarrheas from non-invasive ones in Dhaka, Bangladesh. In that setting, the presence of visible blood in faeces was a reliable indicator of invasive diarrhea. When gross blood could not be seen, occult blood was equivalently reliable, unless a test kit with poor sensitivity was employed. Bardhan *et al*. [13] thus found that FOBT was a valuable test in delineating non-bloody diarrhea in Dhaka, and this was comparable to fecal microscopy outcomes. The aim of this study was to compare the diagnostic efficacy of locally procurable rapid fecal occult blood test (FOBT) kits at identifying invasive infantile diarrhea in northern Ibadan, Nigeria with fecal microscopy as the gold standard.

## Materials and Methods

### Ethical considerations

Ethical approval for this work was obtained from the UI/UCH ethics committee (approval number UI/EC/15/093). Study participants’ guardians provided written informed consent for their participation in the study.

### Fecal occult blood test

A total of four FOBT kits were evaluated in this study alongside the innovator’s product (Cromatest, Spain) (Table 1). Forty-six (46) Fecal specimens from children below 5 years old with diarrhea, being collected as part of a case-control study in our laboratory in Nigeria, were tested according to manufacturers’ instructions for each kit. A sample was either considered negative for FOBT if a single line is spotted on the test strip or positive if two lines (one being the control line) was found on the test strip at the end of the testing procedure. Diagnostic test efficacy was computed as described earlier [14].

**Table 1:**
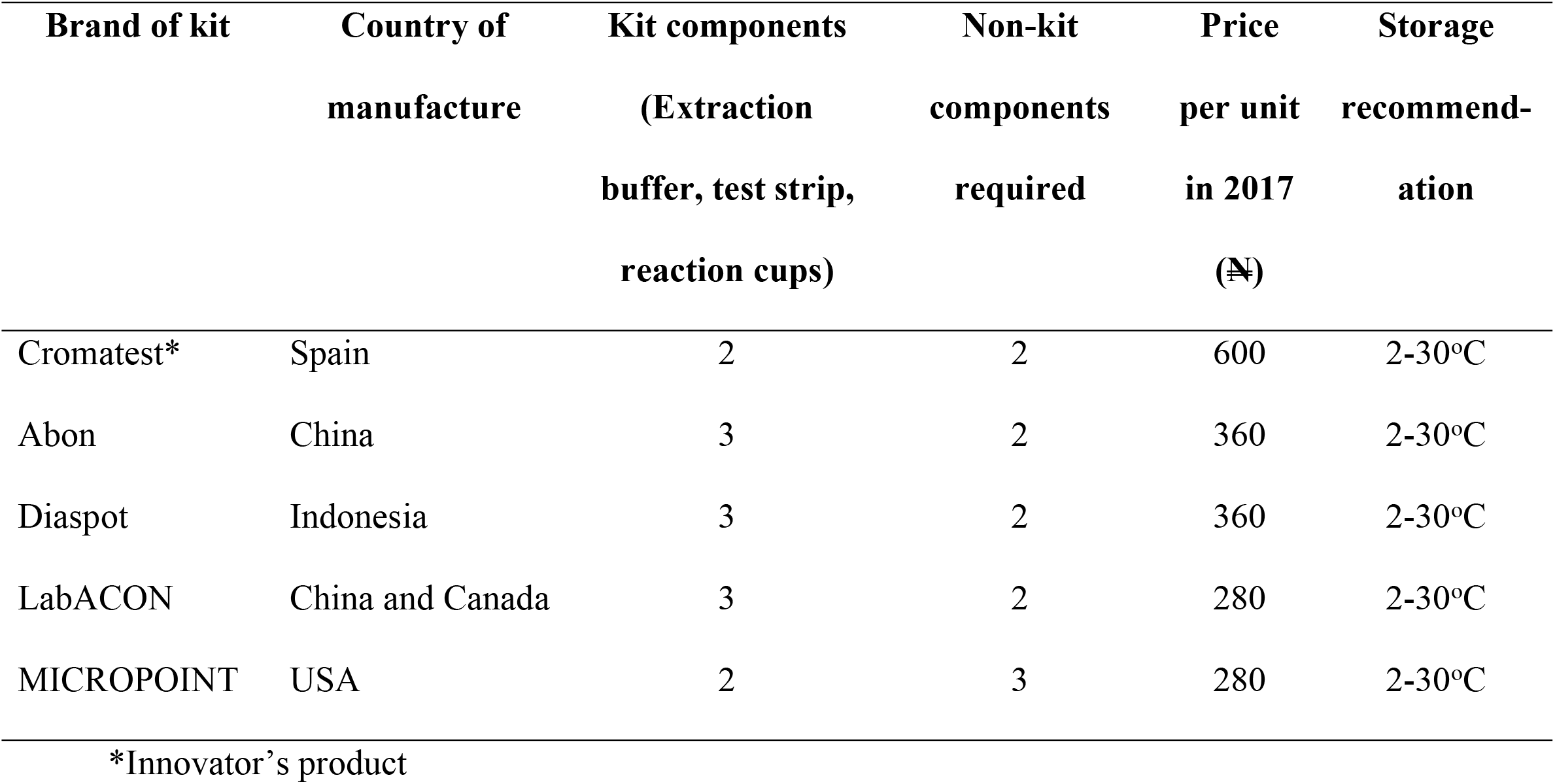
Preliminary information on the five FOBT kits investigated.

### Stool microscopy testing

Fecal microscopy for occult blood (the control assay for the FOBT tests), was done using wet mount method [15] and culture for bacterial pathogens was also performed concomitantly using standard procedures.

### Bacteria culture and biochemical testing

Bacterial culture of stool specimens was performed using standard methods as described by Murray *et al*., 1995 while biochemical identification of strains was carried out using Microbat 12B, 12E and 24E kits. Molecular identification of pathogen subtypes were performed by polymerase chain reaction (PCR), as described previously [16,17]. Briefly, isolate DNA was extracted aseptically using the Wizard Genomic Extraction kit (Promega). Polymerase chain reaction(PCR) for enteropathogenic, enterotoxigenic, enteroinvasive, enteroaggregative and Shiga-toxin-producing *E. coli*, and for *Salmonella enterica* using the methods described earlier [18,19] Identified pathotypes were confirmed by Whole genome sequencing using Illumina platform. Raw reads quality check, assembly, assembly quality check and speciation was done according to Akinlabi *et al*., 2022. Sequence data were submitted to ENA and are available from ENA https://www.ebi.ac.uk/ena/browser/home and Genbank https://www.ncbi.nlm.nih.gov/genbank/ as Bioproject PRJEB8667.

### Data analysis

Data from FOBT assays for each kit were analyzed by comparing outcomes to microscopic data and subsequently estimating sensitivity, selectivity, positive and negative predictive values using standard formulae. Statistical testing was also performed by the Fisher’s Exact Test in EpiInfo Software.

FOBT kit specificity, sensitivity, positive predictive values, and negative predictive values were computed using the following formulae:

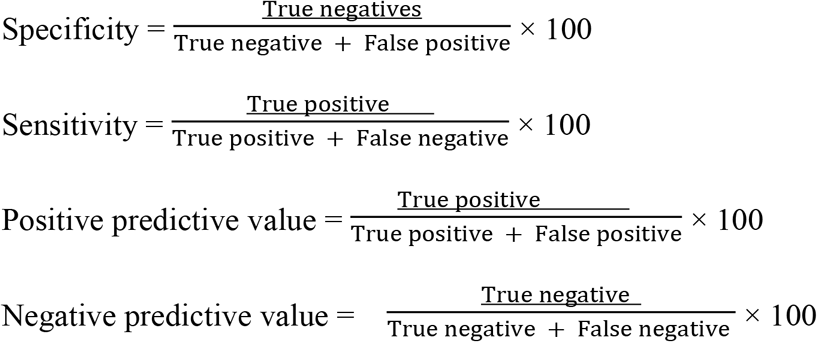

## Results

### Diagnostic efficacy of FOBT kits evaluated

The five kits tested used an easy-to-follow protocol executable in 4-7 minutes using 2-3 kit components and user-supplied sterile swab sticks (Table 1). All tests gave control line results so that none of the test strips used had to be invalided. As shown in Figure 1, a positive result was easy to call for each kit. Twelve of 46 (26.1%) specimens examined gave positive FOBT outcomes with the innovator’s kit (Cromatest), eight of which were also positive by stool microscopy (Table 2). All kits evaluated had comparable specificity with the innovator product (Cromatest), with Diaspot exhibiting slightly higher specificity than the other kits tested, including Cromatest (Table 3). The negative and positive predictive values for the kits were also comparable (Table 3). LabACON and Abon were however insufficiently sensitive compared to the innovator and the other generics kits tested (Table 3).

**Table 2:**
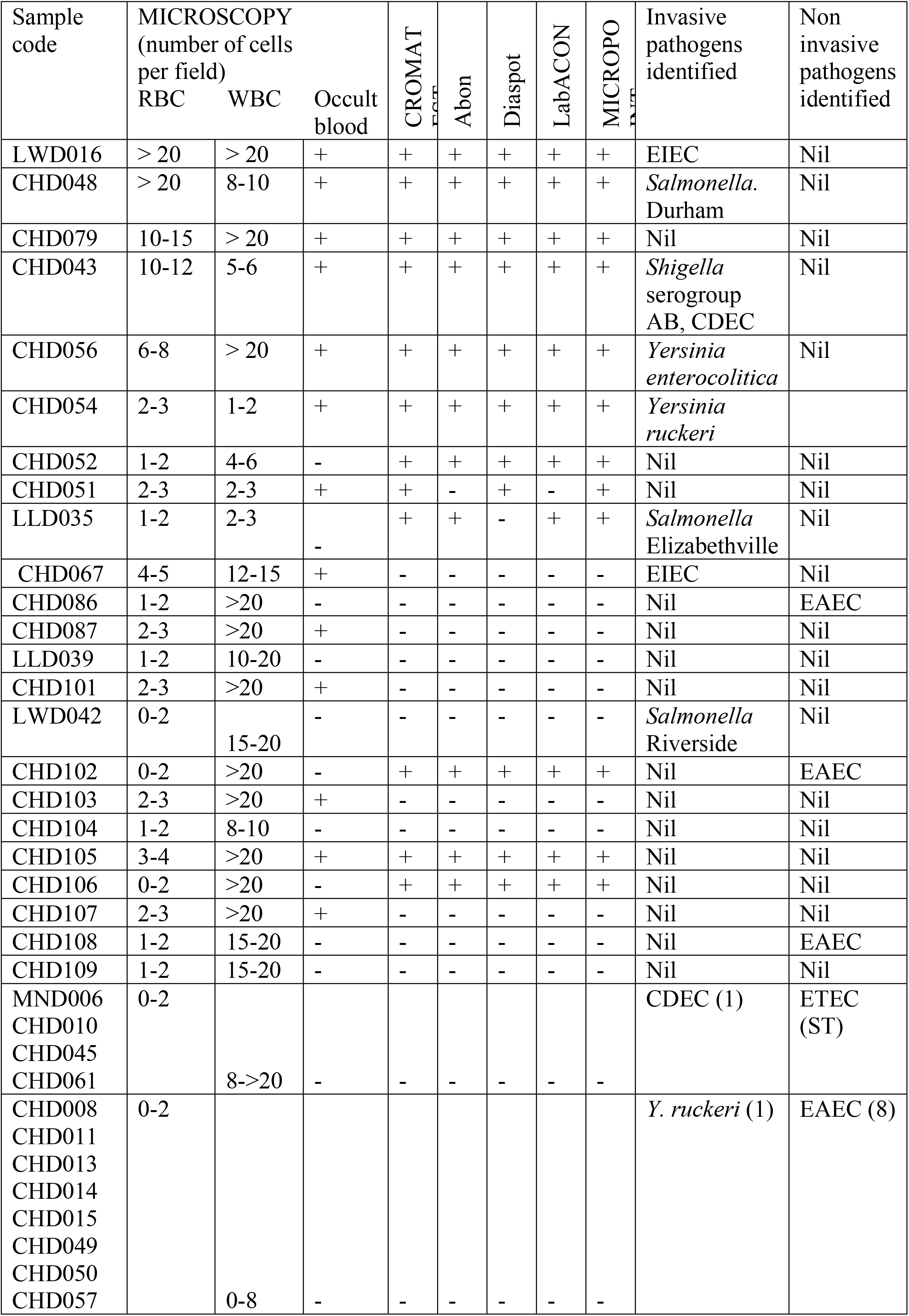
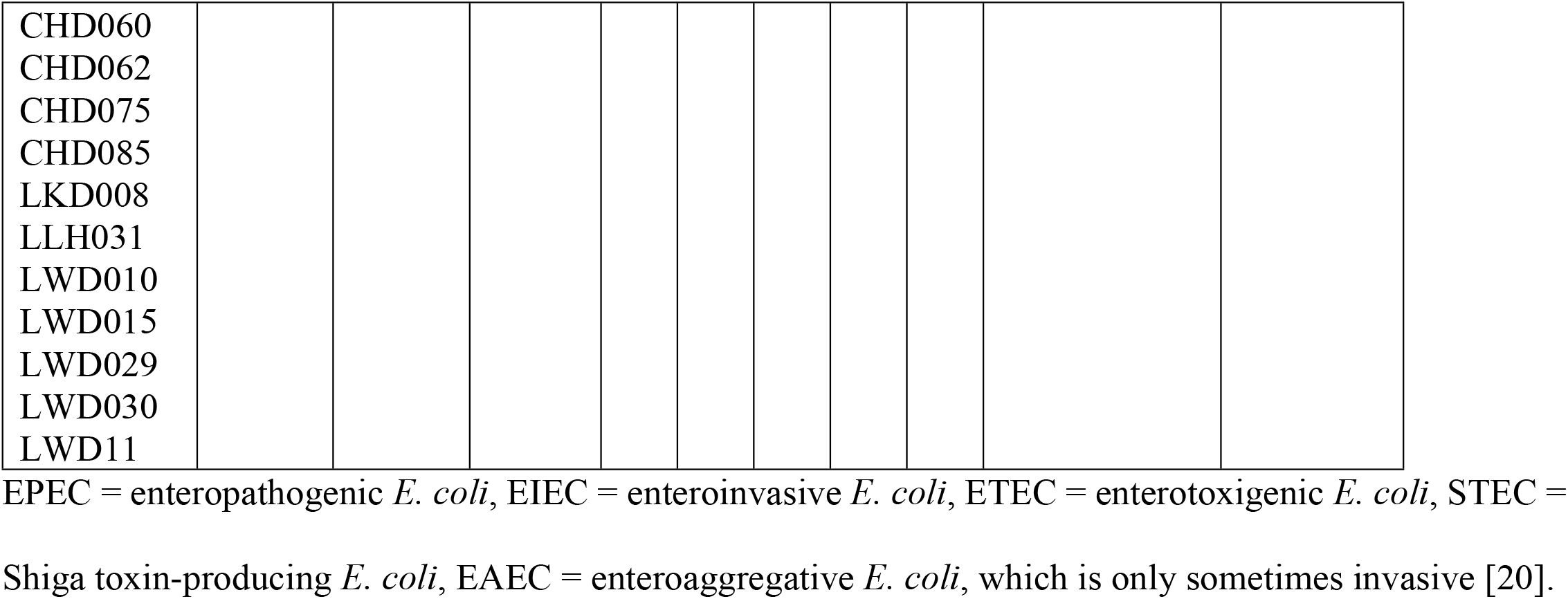
Microscopy, FOBT outcomes and aetiologic agents.

**Table 3:** Diagnostic efficacy of FOBT kits used (presuming that >2 RBCs per field indicates occult blood)

| FOBT Kit | Positives | Negatives | Sensitivity<br>(%) | Specificity<br>(%) | Positive<br>predictive<br>value (%) | Negative<br>predictive<br>value (%) |
| --- | --- | --- | --- | --- | --- | --- |
| Cromatest | 12 | 32 | 61.54 | 87.10 | 66.67 | 84.38 |
| Abon | 11 | 33 | 53.85 | 87.10 | 63.64 | 81.82 |
| Diaspot | 11 | 33 | 61.54 | 90.32 | 72.73 | 84.85 |
| LabACON | 11 | 33 | 53.85 | 87.10 | 63.64 | 81.82 |
| Micropoint | 12 | 32 | 61.54 | 87.10 | 66.67 | 84.38 |

**Figure 1:**
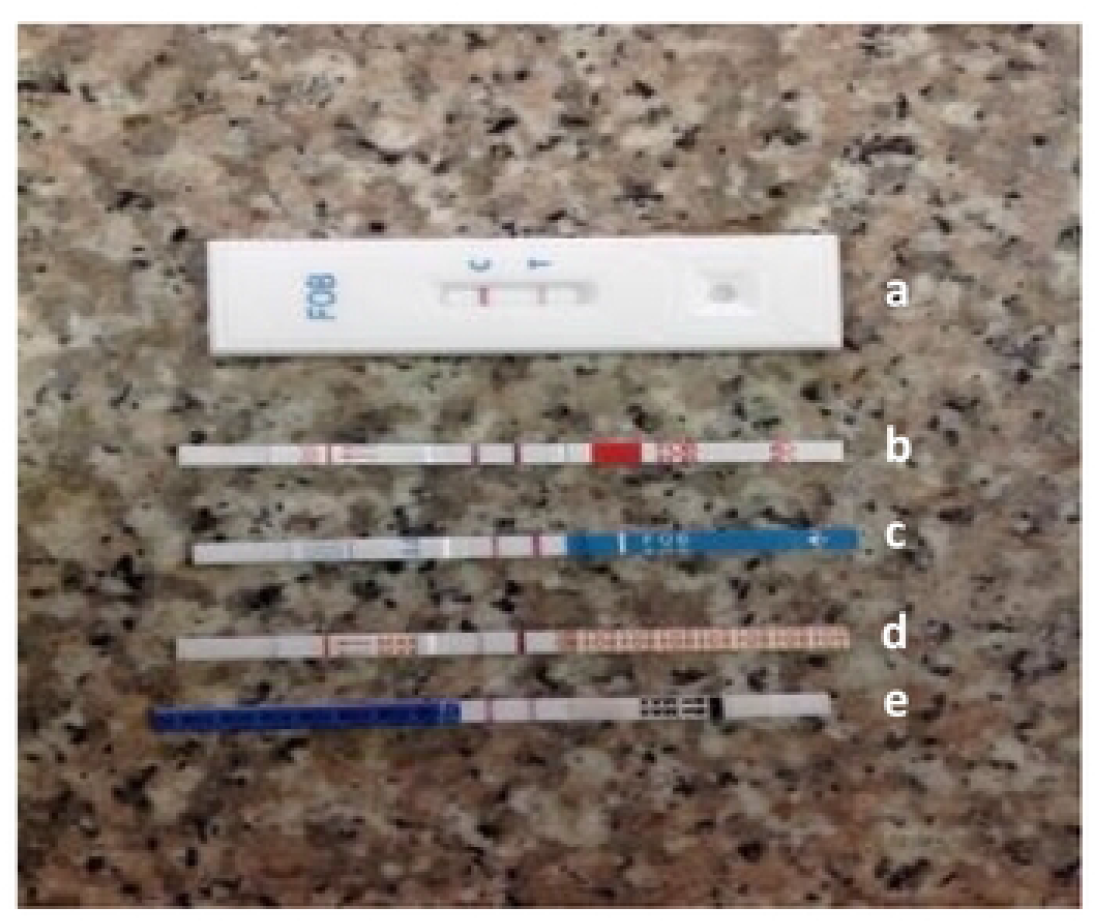
Positive test results for the five FOBT kits tested (a) CROMATEST, (b) Abon, (c) Diaspot, (d) LabACON and (e) Micropoint

Of the bacterial pathogens detected, *Shigella* and enteroinvasive *E. coli* (EIEC), *Salmonella* and *Yersinia*, spp. can produce dysenteric or invasive infections, while enterotoxigenic (ETEC) and enteropathogenic E. coli (EPEC) typically do not. The enteroaggregative *E. coli* (EAEC) pathotype is highly heterogenous and believed to comprise invasive and non-invasive strains however most isolates are believed to be non-invasive. In this study, thirteen of the 46 specimens yielded at least one bacterial pathogen, with five of the children from which these specimens were derived suffering mixed infections. Potentially invasive diarrheal pathogens were detected in nine of the 46 samples tested. Occult blood was detected by microscopy in six of the nine samples from which an invasive bacterial pathogen and six of the cases where no invasive bacterium was detected (p=0.01).

When we compared the performance of FOBT test kits to microscopy, we found that Cromatest, the innovator’s product, had a sensitivity and specificity of 61.5 and 87.1% for occult blood detection respectively, compared to microscopy (Table 3). As shown in Table 3, two of the cheaper products that are readily available in Nigeria (Diaspot and Micropoint) performed as well as the innovator product, with Diaspot showing slightly higher specificity. Abon and LabACON were as specific as Cromatest, but slightly less sensitive.

## Discussion

Unless gross blood or mucus are seen in stools, delineating childhood diarrheas as potentially dystenteric gastrointestinal episodes, which require antimicrobials, is difficult on the basis of clinical signs and symptoms alone. A conventional approach to rapidly detect occult blood in stool by microscopy, which can be performed in under an hour but is tedious, requires trained personnel and uses resources also used for malaria testing in our setting. Culture can identify invasive bacterial pathogens at slightly higher throughput but takes 2-3 days and additionally also requires skilled-labour. We sought to identify a quick, cheap and reliable approach that can be inexpensively implemented in health centres in Nigeria and similar settings irrespective of whether a medical laboratory scientist is available.

The innovator’s FOBT kit is intermittently available in Nigeria and costlier than competitor products. We identified four generic FOBT test kits, which could be procured within Ibadan, Nigeria, a non-coastal city, without an international airport. In our evaluation, all five kits gave similar results and compared reasonably well with the gold standard, expert microscopy.

A large number of potentially invasive bacterial pathogens was detected by culture in the specimens and in six of these nine cases, a positive FOBT result was obtained with any kit. Even though we did not seek protozoal parasites that can elicit blood in stool, significantly higher number of specimens with positive FOBT outcomes had potentially invasive pathogens. Based on our findings, occult blood tests can play a role in identifying diarrheal cases requiring antibacterial therapy when more in-depth lab testing is unavailable. On the average, it takes 4 - 7 minutes to conduct test per specimen using any of the 5 FOBT kits. In 2017-18, each of these kits cost average of N 30 ($ 0.065) at current exchange rates ($1 at N 460.28). The cost and total time taken from sample processing to result demonstrate that FOBT test are fast, cheap, and easy to run.

Our evaluation has some limitations. Because we wished to comparatively evaluate several kits, only a small number of specimens could be screened. The number of specimens that could be evaluated was additionally constrained by Cromatest (the innovator’s product) stock outs. Additionally, we did not seek viral or protozoal pathogens, or *Campylobacter* species and many of these can produce invasive disease. However, even with these limitations, the data appear to suggest that fecal occult blood tests have significant value in delineating children with potentially invasive diarrhea, who should receive antibacterial therapy, and that generic products are functionally equivalent for this purpose to the innovator’s product. Using these kits at the point-of-care in institutions where laboratory testing would normally be unavailable could help to improve patient care and contain antimicrobial resistance.

## Conclusion

FOBT kits are rapid, cost effective and valuable screening processes for quick diagnosis of diarrhea. They are viable alternatives to stool microscopy for paediatric specimens at the point-of-care. Inexpensive, locally available kits perform similarly to the more difficult-to-procure innovator’s product. If used routinely, FOBTs could avoid unnecessary empiric antimicrobial prescription and could also be an early warning indicator for outbreaks due to invasive pathogens.

## Data Availability

Sequence data were submitted to ENA and are available from ENA https://www.ebi.ac.uk/ena/browser/home and Genbank https://www.ncbi.nlm.nih.gov/genbank/ as Bioproject PRJEB8667. All other data are contained in the manuscript.

https://www.ncbi.nlm.nih.gov/bioproject/PRJEB8667/

## Acknowledgement

We thank UNITECH laboratory Services for their assistance in market surveys for FOBT products in Nigeria, Abiodun Oyerinde and Emmanuel Bamidele for technical assistance and A Oladipo Aboderin for helpful discussions.

This work was supported by African Research Leader’s Award MR/L00464X/1 to INO. The award is funded by the UK Medical Research Council (MRC) and the UK Department for International Development (DFID) under the MRC/DFID Concordat agreement and is also part of the EDCTP2 programme supported by the European Union. INO is a Calestous Juma Science Leadership Fellow (Award # INV-036234) supported by the Bill and Melinda Gates Foundation.

## References

1. World Gastroenterology Organisation, Practice Guideline for Acute Diarrhea, 2008.

2. McFarland LV, Elmer GW, McFarland M. Meta-analysis of probiotics for the prevention and treatment of acute pediatric diarrhea. International journal of probiotics and prebiotics. 2006;1(1):63.

3. DuPont HL. Acute infectious diarrhea in immunocompetent adults. N Engl J Med. 2014 Apr 17;370(16):1532–40. doi: 10.1056/NEJMra1301069. PMID: 24738670.

4. Vögtlin J, Stalder H, Hürzeler L, Vischer W, Loosli J, Gyr K, Stalder GA. Modified guaiac test may replace search for fecal leukocytes in acute infectious diarrhea. Lancet. 1983 Nov 19;2(8360):1204. doi: 10.1016/s0140-6736(83)91257-6. PMID: 6139563.

5. Institute for Health Metrics and Evaluation (IHME) Global burden of disease, 2018. Annual deaths from diarrheal diseases differentiated by age categories. Retrieved from https://ourworldindata.org/diarrheal-diseases on 29th January, 2020.

6. Antimicrobial Resistance Collaborators. Global burden of bacterial antimicrobial resistance in 2019: a systematic analysis. Lancet. 2022 Feb 12;399(10325):629–655. doi: 10.1016/S0140-6736(21)02724-0. Epub 2022 Jan 19. Erratum in: Lancet. 2022 Oct 1;400(10358):1102. PMID: 35065702; PMCID: PMC8841637.

7. Angell B, Sanuade O, Adetifa IMO, Okeke IN, Adamu AL, Aliyu MH, Ameh EA, Kyari F, Gadanya MA, Mabayoje DA, Yinka-Ogunleye A, Oni T, Jalo RI, Tsiga-Ahmed FI, Dalglish SL, Abimbola S, Colbourn T, Onwujekwe O, Owoaje ET, Aliyu G, Aliyu SH, Archibong B, Ezeh A, Ihekweazu C, Iliyasu Z, Obaro S, Obadare EB, Okonofua F, Pate M, Salako BL, Zanna FH, Glenn S, Walker A, Ezalarab M, Naghavi M, Abubakar I. Population health outcomes in Nigeria compared with other west African countries, 1998-2019: a systematic analysis for the Global Burden of Disease Study. Lancet. 2022 Mar 19;399(10330):1117–1129. doi: 10.1016/S0140-6736(21)02722-7. Epub 2022 Mar 15. PMID: 35303469; PMCID: PMC8943279.

8. Sharma VK, Vasudeva R, Howden CW. Colorectal cancer screening and surveillance practices by primary care physicians: Results of a national survey. Am J Gastroenterol 2000;95:1551–6.

9. Sharma VK, Corder FA, Raufman JP, et al. Survey of internal medicine residents’ use of the fecal occult blood test and their understanding of colorectal cancer screening and surveillance. AmJ Gastroenterol 2000;95: 2068–73.

10. Hossain MA, Albert MJ. Effect of duration of diarrhea and predictive values of stool leucocytes and red blood cells in the isolation of different serogroups or serotypes of Shigella. Trans R Soc Trop Med Hyg 1991;85:664–6.

11. Ronsmans C, Bennish ML, Wierzba T. Diagnosis and management of dysentery by community health workers. Lancet 1988;2: 552–5.

12. Friedman A, Chan A, Chin LC, Deen A, Hammerschlag G, Lee M, Liddell J, Loh K, Moore E, Ng J, Gibson PR. Use and abuse of fecal occult blood tests in an acute hospital inpatient setting. Internal Medicine Journal 40 (2010) 107–111. 107.

13. Bardhan PK, Beltinger J, Beltinger RW, Hossain A, Mahalanabis D, Gyr K. Screening of patients with acute infectious diarrhea: Evaluation of clinical features, fecal microscopy, and fecal occult bloodtesting. Scand J Gastroenterol 2000;35:54–60.

14. Murray, PR, Baron, EJ, Pfaller, MA, Tenover, FC, Yolken, RH. Manual of Clinical Microbiology, 6th edition. American Society of Microbiology Press, 1995. Washington DC. 1482 p.

15. Arora BB, Maheshwari M, Devgan N, Arora DR. Prevalence of Trichomoniasis, Vaginal Candidiasis, Genital Herpes, Chlamydiasis, and Actinomycosis among Urban and Rural Women of Haryana, India. J Sex Transm Dis. 2014;2014:963812. doi: 10.1155/2014/963812. Epub 2014 Oct 28. PMID: 26316979; PMCID: PMC4437425.

16. Odetoyin BW, Hofmann J, Aboderin AO, Okeke IN. Diarrheagenic Escherichia coliin mother-child Pairs in Ile-Ife, South Western Nigeria. BMC Infect Dis 16, 28 (2015). https://doi.org/10.1186/s12879-016-1365-x

17. Akinlabi OC, Dada RA, Nwoko EQ, Okeke IN. Insufficiency of PCR diagnostics for Detection of Diarrheagenic Escherichia coli in Ibadan, Nigeria. medRxiv 2023.01.06.23284276; doi:https://doi.org/10.1101/2023.01.06.23284276

18. Aranda KR, Fagundes-Neto U, Scaletsky IC. Evaluation of multiplex PCRs for diagnosis of infection with diarrheagenic Escherichia coli and Shigella spp. J Clin Microbiol. 2004 Dec;42(12):5849–53. doi: 10.1128/JCM.42.12.5849-5853.2004. PMID: 15583323; PMCID: PMC535216.

19. Tennant SM, Diallo S, Levy H, Livio S, Sow SO, Tapia M, et al. Identification by PCR of Non-typhoidal Salmonella enterica Serovars Associated with Invasive Infections among Febrile Patients in Mali. PLoS Neglected Tropical Diseases. 2010;4(3):e621.

20. Bouckenooghe AR, DuPont HL, Jiang ZD, Adachi J, Mathewson JJ, Verenkar MP, Rodrigues S, Steffen R. Markers of enteric inflammation in enteroaggregative Escherichia coli diarrhea in travelers. The American journal of tropical medicine and hygiene. 2000 Jun 1;62(6):711–3.

